# Resistance to antibiotics in clinical isolates of *Escherichia coli* producing and non-producing Extended Spectrum Betalactamases (ESBL) obtained from urine cultures in patients with urinary tract infection

**DOI:** 10.1101/2023.04.05.23288193

**Authors:** María Guadalupe Benítez Vergara, María Fernanda Cruz Rosas, Pablo Gustavo Rojas García, Rodolfo García Contreras, Gabriel Martínez Gonzalez, Jorge Angel Almeida Villegas

**Affiliations:** Professional School of Medicine, Nutrition and QFB, University of Ixtlahuaca CUI, Ixtlahuaca, Mexico; Departament of Microbiology and Parasitology, Medicin School, National Autonomous University of Mexico, Mexico City, Mexico; Health Research Institute (QFB-INIES), University of Ixtlahuaca CUI, Ixtlahuaca, Mexico

**Keywords:** Resistance, antibiotics, *E coli*, sensitivity, ESBL

## Abstract

**Objective:** Identify resistance patterns against various antibiotics in *Escherichia coli* producers and non-producers of extended spectrum beta-lactamases in urinary infections in a population of the Toluca Valley, Mexico

**Introduction:** *Escherichia coli* is a bacterium that is part of the normal biota of the human being, but under certain conditions it can produce diseases such as gastrointestinal and urinary tract infections for which is the main responsible. *Enterobacteriaceae* such as *Escherichia* are extended spectrum betalactamases (ESBL) producers, which makes their treatment difficult due to a high rate of resistance to antibiotics.

**Methods:** 155 samples were collected from patients with suspected urinary tract infection without exclusion criteria such as age or gender. Automated equipment was used for the identification of the etiological agent and sensitivity tests. For the determination of ESBL, the double disc technique was used.

**Results:** 35 strains of *Escherichia coli* were obtained, of which 45.72 % have ESBL, these strains show 100 % resistance to betalactams, as well as high resistance to quinolones and tetracycline ranging from 70 to 100 %. On the other hand, the resistance shown by non-ESBL producing strains is variable both for betalactams and for other antibiotics.

**Conclusion:** Treatment for urinary tract infections has become increasily difficult due to the high rate of resistance to antibiotics, mainly ESBL-producing strains such as the *E coli* strains in the present study. Where high rates of resistance to different antibiotics used in clinical practice are shown.

## Introduction

*Escherichia coli* is commonly found in the normal microbiota in the human gastrointestinal tract and is intricately involved in the lives of humans^1^. *Escherichia coli* typically colonizes the gastrointestinal tract of human infants within a few hours after birth. Usually, *E. coli* and its human host coexist in good health and with mutual benefit for decades. These commensal *E. coli* strains rarely cause disease except in immunocompromised hosts or where the normal gastrointestinal barriers are breached as in peritonitis, for example^2^. In addition, six pathotypes of diarrheagenic *E. coli* are recognized, each with distinct phenotypic and genetic traits^3^. Diarrhoeagenic *Escherichia coli* is categorized into the following six pathotypes: enteropathogenic *E. coli* (EPEC), enterotoxigenic *E. coli* (ETEC), enterohaemorrhagic *E. coli* (EHEC), enteroinvasive *E. coli* (EIEC), diffusely adherent *E. coli* (DAEC), and enteroaggregative *E. coli* (EAEC). Other diarrhoeagenic *E. coli* pathotypes have been proposed, such as cell detaching *E. coli* (CDEC); however, their significance remains uncertain^4^.

Some *E. coli* strains can cause a wide variety of intestinal and extra intestinal diseases, such as diarrhea, urinary tract infections, septicemia, and neonatal meningitis. Phylogenetic analyses have shown that *E. coli* strains fall into four main phylogenetic groups (A, B1, B2, and D) and that virulent extra-intestinal strains belong mainly to group B2 and, to a lesser extent, to group D whereas most commensal strains belong to group A.^5^

Urinary Tract Infections (UTIs) are among the most common bacterial infections, affecting 150 million people worldwide each year. Although both men and women may become infected, UTIs are traditionally considered a disease of women, among whom 50 % will be affected during their lifespan.^6^ Symptomatic urinary tract infection in a healthy woman is a complex event. It is initiated when potential urinary pathogens from the bowel, or in some cases from the vagina (as a result of direct inoculation during sexual activity), colonize the periurethral mucosa and ascend through the urethra to the bladder and in some cases through the ureter to the kidney.^7^

UTI is diagnosed using a combination of urinary symptoms and urine culture demonstrating numbers of a known uropathogen above a given threshold (usually defined as >1,000 CFU/ml of urine, but thresholds as low as 100 CFU/ml and as high as 100,000 CFU/ml are also used). However, urinary symptoms and bacteriuria frequently occur independently of each other: ∼20% of women presenting with ‘classic’ UTI symptoms have negative results in urine cultures. Significant numbers of bacteria are often found in the urine of otherwise healthy, asymptomatic individuals.^8^

*Escherichia coli* is the most common cause of UTIs, with both symptomatic and asymptomatic infections most often being associated with specific uropathogenic *E. coli* (UPEC) sublineages.^9^ Four main UPEC phylogroups (A, B1, B2, and D) have been identified on the basis of the occurrence of genomic Pathogenicity Islands (PAI) and the expression of virulence factors, such as adhesins, toxins, surface polysaccharides, flagella, and iron-acquisition systems. Usually, many of these virulence factors are required for UPEC to cause UTI. During UTIs, UPEC pathogenesis includes: (a) UPEC colonization of the periurethral and vaginal areas with colonization of the urethra; (b) ascending into the bladder lumen and growth as plantktonic cells in urine; (c) adherence to the surface and interaction with the bladder epithelium defense system; (d) biofilm formation; (e) invasion and replication by forming bladder Intracellular Bacterial Communities (IBCs) where quiescent intracellular reservoirs (QIRs) form and reside in the underlying urothelium; (f) kidney colonization and host tissue damage with increased risk for bacteremia/septicemia ^10^.

UPEC strains have been shown to grow better in urine than nonpathogenic strains; in fact, UPEC strains grow extraordinarily fast in urine, and rapid growth has been proposed as a virulence factor. Urine composition is likely to be crucial for maintaining a healthy genitourinary microbiome or encouraging UPEC growth. Age-related changes or fluctuations in urine composition may account for increased susceptibility to infection^11^.

UTIs are routinely treated with antibiotic therapy, including trimethroprim–sulfamethoxazole (TMP–SMX), ciprofloxacin^12^, aminoglycosides, oral cephalosporins and nitrofurantoin^13^.

The therapeutic options for UTI caused by *E. coli* have been progressively reduced due to the increasingly frequent presence of extended-spectrum betalactamases (ESBLs), which are plasmid-mediated enzymes with the ability to hydrolyze penicillins, oxymino-cephalosporins, extended spectrum cephalosporins and aztreonam^14^. Bacterial infections are still a leading cause of death, and options for treating these infections are declining, due to the increase in bacteria resistant to antibiotics. In fact, several bacterial strains are currently resistant to virtually all known antibiotics.^15^

## Methods

In the present study, 155 urine cultures were performed in patients with clinical suspicion of urinary tract infection, without exclusion criteria such as gender or age. In a town in the center of the Toluca Valley, Mexico.

Of the 155 urine cultures performed, the distribution was; 107 people of the female gender representing 69 % of the study population and 48 men representing 31 %. The age ranges in the lower limit is a 1-year-old female patient and for the higher limit is a 91-year-old patient, with an average of 30 years.

For the study, 100 ml of the first urine of the day were collected with the necessary hygiene conditions for the collection of the specimen.

An aliquot was taken with a calibrated handle, and blood agar was inoculated to perform the colony count and chromogenic agar. Both Petri dishes were incubated for 24 hours at 37°C.

An isolated colony was taken from the chromogenic agar medium and suspended in the diluent medium, which was placed in the plates of the automated equipment, adjusting to a 0.5 McFarland scale.

The identification of the etiological agent as well as the sensitivity tests were performed using an automated method using the walkaway SI 96 from Beckman coulter equipment. For the detection of spread spectrum betalactamases, the double disc method was used.^16^

For the statistical study, a Shapiro Wilk normality test, a non-parametric Spearman correlation, frequency distribution and one-way ANOVA were performed.

## Results and Discussion

Of the urine cultures obtained and performed, 80 of them were positive for the development of microorganisms, which represents 51.6 %, on the other hand, 48.4 % representing 75 cultures that did not present microbial development.

Of the 51.6 % that had microorganism growth, 47.5 % represented bacteria other than *E coli*, 8.75 % corresponded to yeasts and 43.75 % represented *E coli*, which is equivalent to 35 cultures. Of the 35 isolated from *Escherichia coli*. 16 strains show production of extended spectrum betalactamases (ESBLs) equivalent to 45.72 %, and 19 isolates that do not show ESBLs production. Table 1 shows the susceptibility patterns to different antibiotics used for non-ESBLs-producing strains of *E coli*.

**Table 1.** Antibiotic sensitivity patterns as resistant and sensitive for *E coli* not producers of ESBL.

| Antibiotics | Sensitivity pattern |  |
| --- | --- | --- |
|  | Resistant | Sensitive |
| Amikacin | 0 % n=0 | 100 % n=19 |
| Amoxicillin /Clavulanic Acid | 15.79 % n=3 | 84.21 % n=16 |
| Ampicillin | 73.68 % n=14 | 26.32 % n=5 |
| Ampicillin /Sulbactam | 42.11 % n=8 | 57.89 % n=11 |
| Cefazolin | 26.32 % n=5 | 73.68 % n=14 |
| Cefepime | 21.06 % n=4 | 78.94 % n=15 |
| Cefotaxime | 15.79 % n=3 | 84.21 % n=16 |
| Ceftazidime | 21.06 % n=4 | 78.94 % n=15 |
| Ceftriaxone | 26.32 % n=5 | 73.68 % n=14 |
| Cefuroxime | 26.32 % n=5 | 73.68 % n=14 |
| Ciprofloxacin | 26.32 % n=5 | 73.68 % n=14 |
| Ertapenem | 0 % n=0 | 100 % n=19 |
| Gentamicin | 15.79 % n=3 | 84.21 % n=16 |
| Imipenem | 0 % n=0 | 100 % n=19 |
| Levofloxacin | 36.85 % n=7 | 63.15 % n=12 |
| Meropenem | 0 % n=0 | 100 % n=19 |
| Piperacillin/ Tazobactam | 0 % n=0 | 100 % n=19 |
| Tetracycline | 68.42 % n=13 | 31.58 % n=6 |
| Tigecycline | 0 % n=0 | 100 % n=19 |
| Tobramycin | 31.58 % n=6 | 68.42 % n=13 |
| Trimethoprim/Sulfamethoxazole | 47.37 % n=9 | 52.63 % n=10 |

It was found that the 19 isolates were 100 % sensitive to the aminoglycoside amikacin, but not to gentamicin, which shows 15.79 % resistance. In carbapenems (imipenem, ertapenem and meropenem), as well as piperacillin with tazobactam there is 100 % sensitivity. While 68.42 % equivalent to 13 strains were resistant to tetracycline, but 100 % sensitive to tigecycline. The folate synthesis inhibitors trimethoprim/sulfamtoxazole rank third in resistance with resistance in nine isolates. On the other hand, the antibiotics that showed greater sensitivity than carbapenems are gentamicin, amoxicillin with clavulanic acid, and cefotaxime. The rest of the antibiotics used show patterns of resistance and variable sensitivity as shown in Figure 1.

**Figure 1.**
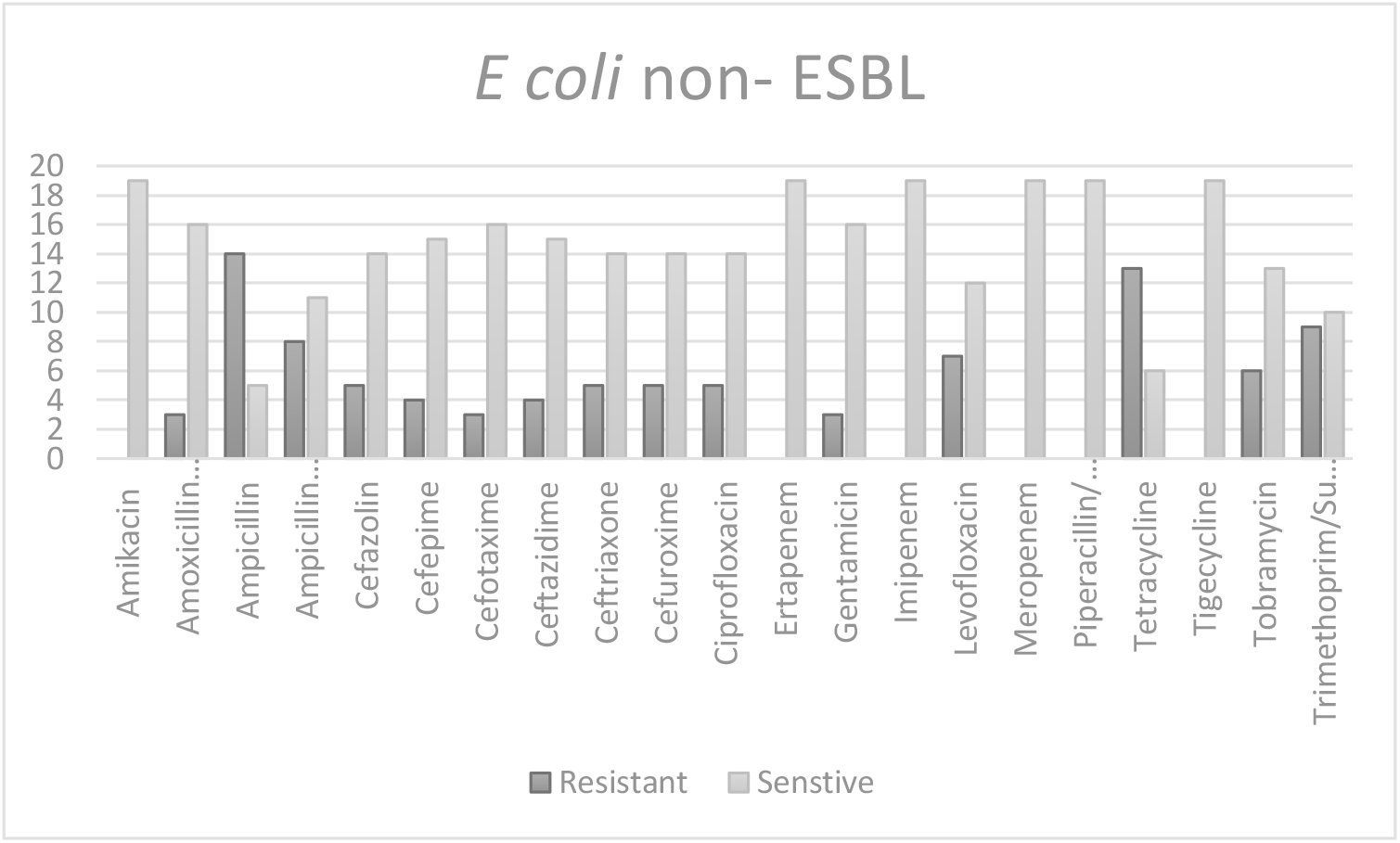
Resistance and sensitivity patterns of the *E. coli* isolates, in black the resistant isolates, and gray strains with sensitivity to the antibiotic.

Table 2 shows the patterns of resistance and sensitivity to different antibiotics in isolated strains of *E coli* producing extended spectrum beta-lactamases. The same antibiotics that were used with non-productive E *coli* strains of ESBL, in order to evaluate the differences in the sensitivity patterns.

**Table 2.** The following table shows the antibiotic sensitivity patterns as resistant, intermediate sensitivity and sensitive for *E coli* producers of ESBL.

| Antibiotics | Sensitivity pattern |  |
| --- | --- | --- |
|  | Resistant | Sensitive |
| Amikacin | 0 % n=0 | 100 % n=16 |
| Amoxicillin /Clavulanic Acid | 37.5 % n=6 | 62.5 % n=10 |
| Ampicillin | 100 % n=16 | 0 % n=0 |
| Ampicillin /Sulbactam | 81.25 % n=13 | 18.75 % n=3 |
| Cefazolin | 100 % n=16 | 0 % n=0 |
| Cefepime | 100 % n=16 | 0 % n=0 |
| Cefotaxime | 100 % n=16 | 0 % n=0 |
| Ceftazidime | 100 % n=16 | 0 % n=0 |
| Ceftriaxone | 100 % n=16 | 0 % n=0 |
| Cefuroxime | 100 % n=16 | 0 % n=0 |
| Ciprofloxacin | 81.25 % n=13 | 18.75 % n=3 |
| Ertapenem | 0 % n=0 | 100 % n=16 |
| Gentamicin | 62.5 % n=10 | 37.5 % n=6 |
| Imipenem | 0 % n=0 | 100 % n=16 |
| Levofloxacin | 87.5 % n=14 | 12.5 % n=2 |
| Meropenem | 0 % n=0 | 100 % n=16 |
| Piperacillin/ Tazobactam | 31.25 % n=5 | 68.75 % n=11 |
| Tetracycline | 93.75 % n=15 | 6.25 % n=1 |
| Tigecycline | 15.2 % n=2 | 87.5 % n=14 |
| Tobramycin | 62.5 % n=10 | 37.5 % n=6 |
| Trimethoprim/Sulfamethoxazole | 75 % n=12 | 25 % n=4 |

Extended-spectrum β-lactamase (ESBL)-producing Gram-negative pathogens are a major cause of resistance to expanded-spectrum β-lactam antibiotics.^17^ The strains show marked 100 % resistance to second generation and third-generation cephalosporins that are broad-spectrum antibiotics, as well as ampicillin with the same percentage, while for tetracycline 93.75 % resistance is shown.

Carbapenems and amikacin show full sensitivity. Other antibiotics such as quinolones, sulfa and other beta-lactams show variable resistance as show in Figure 2.

**Figure 2.**
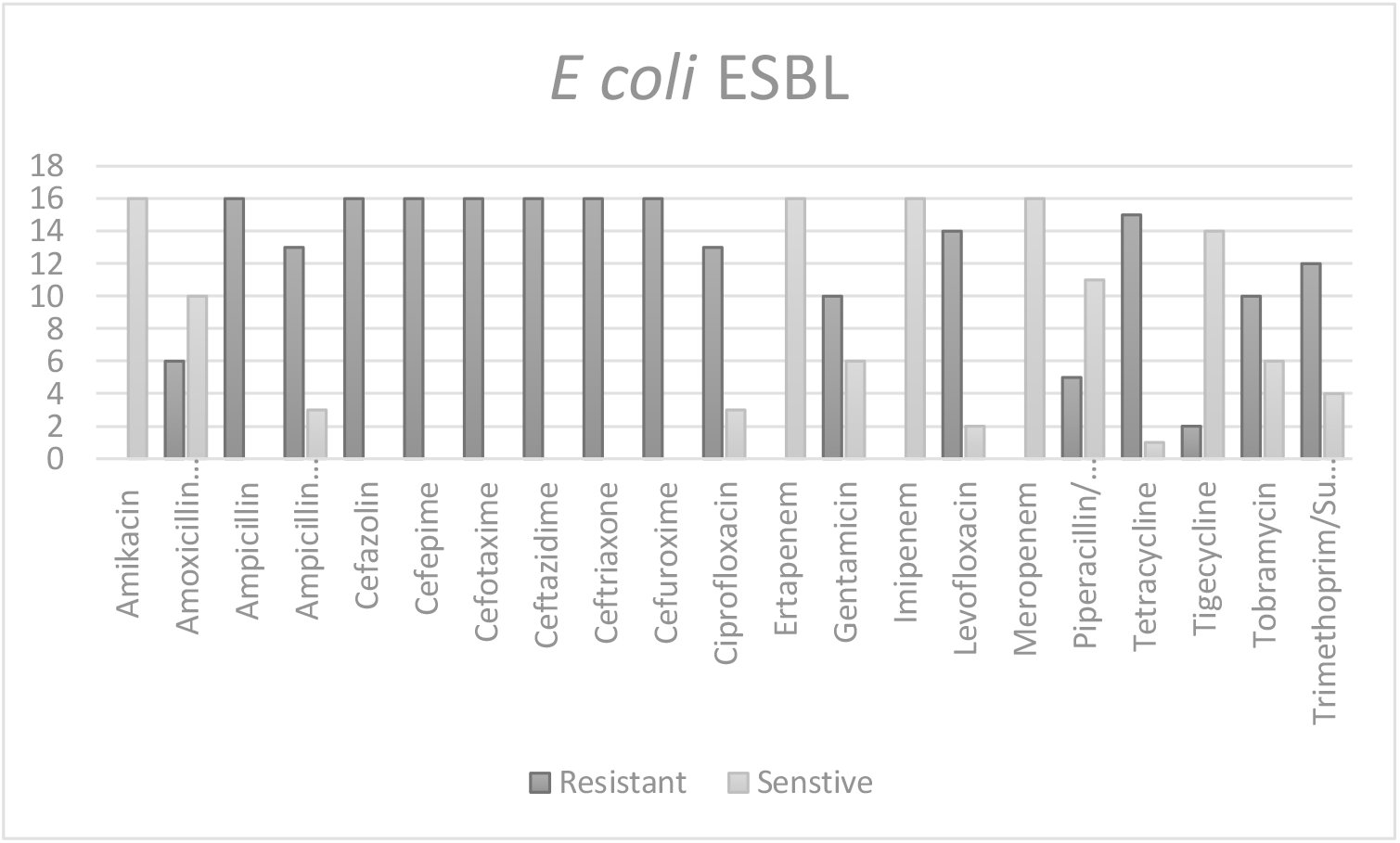
The following graph shows in detail the resistance and sensitivity patterns, in blue the resistant isolates, red represents the strains with intermediate sensitivity and green strains with sensitivity to the antibiotic.

Extended-spectrum beta-lactamases allow enterobacteria to avoid the effect of beta-lactam antibiotics such as first-, second-, third-, fourth-generation cephalosporins, and penicillin derivatives, as this is reflected in the ampicillin sensitivity patterns, that show 100 % to resistance.

In the case of cephalosporins, the sensitivity in non-ESBL-producing strains is 73 to 84 %, and the sensitivity among the different studied, while for the ESBL-producing strains, the sensitivity pattern is 0 %. For the resistance in the non-ESBL-producing it can be caused by other types of mechanisms such as Amp C-type beta-lactamases which mainly affect first and even third generation cephalosporins.^18^

For mixtures of beta-lactams with beta-lactamase inhibitors in non-ESBL-producing isolates, a sensitivity of 84.21 % is achieved for amoxicillin with clavulanic acid and 57.89 % for ampicillin with sulbactam, while for the latter case, ampicillin alone is only 26.31 %. In the case of ESBL-producing strains, the sensitivity shown in ampicillin with sulbactam is only 18.75 %. ESBL enzymes are inhibited by the so-called ‘classical’ β-lactamase inhibitors such as clavulanic acid, sulbactam and tazobactam.^19^ Therefore, resistance to beta-lactams with beta-lactamase inhibitors is lower in ESBL-producing strains as opposed to non-producing ones, since the latter may have the presence of Amp C-type beta-lactamases.

Another factor to consider is the high resistance to tetracyclines and fluoroquinolones, suggesting that the strains carring the Extended-spectrum beta-lactamases extent also have resistance genes towards these antibiotics which is shown for example with 93.75 % resistance to tetracycline in ESBL-producing isolates and only 68.42 % in non-ESBL-producing strains. ^20^

In the case of ciprofloxacin, the non-ESBL strains have a resistance pattern of 26.31 %, while for the ESBL strains the resistance is 81.25 % the patterns of resistance to levofloxacin are greater than those of ciprofloxacin, but this is a misleading result, since being chemically similar in structure, the same resistance mechanisms apply to it. On the other hand, it is known that the presence of ESBL determines resistance to fluoroquinolones through cross-resistance.^21,22^

For the statistical study, a Shapiro Wilk normality test was performed to determine if the data has a normal distribution, for all the tests, since they did not follow a normal distribution, a non-parametric Spearman distribution test was performed and it was as I work with the data. One of the main relationships is whether the frequency of infections, that is, having one or more infections per year conditioned the appearance of ESBL-producing E. coli, when there are less than two infections per year there is no correlation associated with the presence of ESBL.. On the other hand, when there are more than two infections per year, the presence of ESBL-producing E. coli p ≥0.281 has been demonstrated. Another of the points that was determined was the correlation between age and the presence of E. coli ESBL, since it did not follow a normal distribution, a non-parametric Spearman correlation was performed, where there is no direct correlation between age and the ESBL presence. However, if a frequency distribution is performed for the segmentation by age with an amplitude size of 13 and with a one-way ANOVA of the groups resulting from the segmentation, it is found that the group from 27 to 44 years is the one with greater production. Of presents ESBL (p ≥ 0.545). For sex as a step with age, a normal distribution pattern is not followed, but when performing a one-way ANOVA, a clear tendency is observed for the female gender to have ESBL-producing E. coli (p ≥ 0.318). A last correlation that was made was in the strains that have the presence of ESBL and its cross-resistance to fluoroquinolones where there is a high correlation for ciprofloxacin (p ≥ 0.842) and levofloxacin (p ≥ 0.749). This supports the mechanisms described in the literature.

## Conclusions

Urinary tract infections are a type of illness that mainly affects women at an early age, as evidenced by the number of samples obtained: 107 from women and 48 from men. The main bacterium isolated from these infections is *Escherichia coli*, which represents a serious threat to health, since enterobacteria produce extended-spectrum beta-lactamases, which makes therapeutic options difficult, both for betalactams derived from penicillins and cephalosporins. The high presence of ESBL conditioned by age, sex and number of infections also influences the resistance of other families of antibiotics. Clinical isolates have a 100 % resistance percentage for all cephalosporins and ampicillin in the case of ESBL producers, as well as a notable transfer of resistance to quinolones and tetracyclines. The treatment of these conditions begins to be a limitation, due to the resistance of the strains to different antimicrobials, such as *E. coli* in this study.

## Data Availability

Yes

## Ethical approval

The following study was approved by the ethical committee of the research division of the Faculty of Medicine UNAM.

No permission of the patients was required for the obtention of the clinical isolates since they are no human material and are routinely recovered from patients cultures.

## Interest Conflict

The authors declare that they have no conflict of interest.

### Financing

No funding was received to carry out this work.

## Acknowledgment

We thank all the institutions that allowed us to carry out this research work.

